# Healthy diets, lifestyle changes and wellbeing during and after lockdown: Longitudinal evidence from the West Midlands

**DOI:** 10.1101/2021.03.19.21253951

**Authors:** Thijs van Rens, Petra Hanson, Oyinlola Oyebode, Lukasz Walasek, Thomas M. Barber, Redzo Mujcic, Lena Al-Khudairy

## Abstract

**Background:** ‘Lockdowns’ to control the spread of COVID-19 in the UK have affected many aspects of life, with concerns that they may have adversely affected diets. We aimed to examine (i) the effect of living in lockdown on fruit and vegetable consumption; (ii) whether any population subgroup was particularly adversely affected; (iii) the barriers and facilitators to a healthy diet in lockdown; and (iv) the effect of lockdown on secondary outcomes such as weight and mental wellbeing.

**Methods:** We conducted a mixed-method longitudinal study, involving an online survey of 1003 adults in the West Midlands, UK, 494 of whom were surveyed at two different points in time. Our first time point (T0: May 2020) was during stringent COVID-19 lockdown and the second (T1: September 2020) during a period of more relaxed restrictions. The survey included detailed quantitative questions about fruit and vegetable consumption; questions on physical activity, socio-demographic characteristics, BMI and wellbeing; and qualitative data collection about the reasons behind reported changes.

**Results:** We find no evidence for respondents decreasing their fruit and vegetable consumption during lockdown compared to afterwards. If anything, consumption of fruit and vegetables increased by about half a portion daily among women, particularly among those who normally have a long commute. These findings combined with a significant increase in physical activity, suggest that behaviours were healthier during lockdown, consistent with higher self-reported health compared with afterwards. However, there was a marked deterioration in wellbeing during lockdown, and on average participants self-reported being heavier during this period as well. Our qualitative data suggested that an abundance of resources supported higher fruit and vegetable consumption during lockdown, for instance, participants had more time, while access issues were one barrier to consumption.

**Conclusions:** Our results are reassuring for those concerned that lockdowns may have adversely affect diets. They may point to the impact of commuting on diet, particularly for women, and intervening to reduce commuting times may be one way to improve population diets. Our study adds longitudinal evidence to a growing body of literature on the adverse effect of lockdown on mental health.

## BACKGROUND

Covid-19 first appeared in Wuhan, China in December 2019 and within three months spread around the world causing a pandemic, officially declared on March 11, 2020(1). There is much variety in the clinical presentation of Covid-19 (ranging from asymptomatic to death), and clear risk factors regarding susceptibility to and severity of infection, including age, BMI and underlying health conditions including respiratory disease and Diabetes Mellitus (2, 3).

In response to the pandemic, several governments implemented national ‘lockdowns’, in which travel and social activities were restricted, in an attempt to control viral spread. The UK Prime Minister announced a national lockdown on the 23^rd^ of March 2020 in which people were confined to their homes, unless they were deemed critical workers, except for exercise once daily and other limited exceptions (4).

The full implications of the ‘lockdown’ on the health and wellbeing of the population are not yet known. However, there have been concerns that lockdown has adversely affected physical and mental health, as well as health behaviours, possibly due to stress, changes in financial circumstances and lack of access to products and services. Emerging international literature, based predominantly on cross-sectional evidence, has partially supported these fears (5-12).

In the UK, the food sector experienced strained supply and distribution resulting from unusual shopping activities such as panic-buying and stock-piling, affecting major retailers, online stores and local supermarkets (13). In response, food retailers decreased their range of products and focused on products that were in greater demand such as long-life milk, pasta and rice (14).

Further concern has been expressed that unintended consequences of lockdowns may disproportionately affect some population groups, including those who are already disadvantaged for example those with overweight or obesity, as reported in this UK cross-sectional study (15). Further international evidence suggests polarisation in attitudes towards food consumption during the lockdown, such that some people become unhealthier in their lifestyle, whereby others adopt pro-healthy attitudes. For example, evidence from Poland (16), Spain (17, 18) and Qatar (19), indicates that at least some individuals’ diets benefit from the imposition of a national lockdown.

This longitudinal mixed-methods study is focussed on the effect of the UK lockdown on fruit and vegetable consumption. The World Health Organisation recommends that we consume 400g or 5 portions of 80g of fruits and vegetables daily. As a major source of fibre, consumption of fruits and vegetables is associated with multiple health benefits, including improved insulin sensitivity, reduced cardiovascular risk, reduced risk for colorectal carcinoma, improved gastrointestinal function and reduced mortality (20). Fruit and vegetables are also a source of vitamins and minerals. Fruit and vegetable consumption has been shown to be associated with both physical health (all-cause, cancer and cardiovascular disease mortality) (21) and subjective wellbeing (22, 23).

In this study, we aimed to examine the following research questions:

1. Was fruit and vegetable consumption during the lockdown lower than during normal times in the West Midlands, UK?
2. Was fruit and vegetable consumption affected in different ways in different population subgroups defined by individual characteristics? Here, we have a specific interest in whether more vulnerable or less healthy subgroups were more likely to reduce their fruit and vegetable consumption during lockdown than other groups.
3. Were wellbeing, physical activity, behaviour, self-reported weight, and self-rated health different during lockdown compared with other times?
4. What were the barriers and facilitators of fruit and vegetable consumption during lockdown?

## METHODS

We used an observational study design consisting of two online surveys. Participants were recruited through the Prolific platform for online research studies (24) and were selected based on their residency of the West Midlands. This region is interesting as a case study because of its socio-economically diverse populations, particularly in terms of age and income levels (25). We developed an online questionnaire using Qualtrics to capture dietary intake of fruit and vegetables (our primary outcome), self-reported height and weight, physical activity, and measures of physical health, mental health and wellbeing (our secondary outcomes) as well as socio-demographic characteristics. We collected baseline measures (T0) in May 2020, and follow up measures (T1) in September 2020. These dates were chosen to correspond to “during lockdown’’ and “post lockdown” periods. Importantly, the same participants completed both baseline (T0) and follow up (T1) questionnaires, so that we were able to measure within-person changes. This approach helps to overcome a common issue with cross-sectional surveys and studies on health behaviours, where an observed relationship may be spurious due to (perhaps unobserved) differences in the composition of the sample in different time periods. We received ethical approval under the DR@W2 agreement from the Humanities and Social Sciences Research Ethics Committee (HSSREC) at the University of Warwick (reference number 168/19-20).

### Outcomes

We collected information on socio-demographic characteristics, living and working arrangements, mental health, wellbeing, exercise, dietary intake of fruit and vegetables, and self-reported height, weight and health. We administered the Office for National Statistics (ONS) wellbeing tool to measure subjective wellbeing (26) and the short version of the Warwick-Edinburgh Mental Wellbeing scale (SWEMWBS) to measure mental wellbeing (27). To measure fruit and vegetable consumption, we used questions from the Health Survey for England, (HSE) slightly modified for the format of an online survey (28). Consumption of fruit and vegetables is aggregated according to dietary recommendations, as described in the HSE documentation. For broad categories of vegetables (salad, pulses, other vegetables, dishes made mostly from vegetables) and fruit (juice, fresh fruit, dried, frozen, tinned fruit, dishes made mostly from fruit), the questions asked participants whether they consumed any of these during the last 24 hours. If participants answered “yes”, then we also asked how many (bowls, pieces, tablespoons, glasses) they consumed for each type of fruit/vegetable within these categories. Since this was an online survey, we limited the possible answers to the 4 to 6 most common types of fruit or vegetables in each category and added an “other” option, see the questionnaire in supplement 1 for more detail. We then converted the natural quantities reported to the equivalent standard 80g portions, based on the 5-a-day approach according to the HSE documentation, and aggregated over types within categories and over categories. The exact questions may be found in the questionnaires in supplement 1. At follow-up we added free-text questions about the reasons for reported changes in fruit and vegetable consumptions and weight compared to during lockdown.

### Analysis

#### Quantitative outcomes

Descriptive data for continuous measures are presented as means and standard deviations. Descriptive data for categorical measures are presented as fractions. We tabulated changes in outcomes “after” versus “during” lockdown using information collected from our two surveys (T1 and T0, respectively), and “during” versus “before” lockdown using retrospective questions where it was possible to ask these. In all cases, changes refer to outcomes reported by the same individual at different points in time, so that our results reflect true changes in outcomes and are not driven by changes in the composition of the sample.For our main outcomes (fruit and vegetable consumption, and mental health as a secondary outcome), we also cross-tabulate these changes with individual characteristics to explore heterogeneity in the changes in outcomes across subgroups along observable dimensions. The quantitative analysis was executed using the statistical software package Stata.

#### Qualitative outcomes

We applied thematic analysis (29) to analyse the qualitative data from these questions. This analysis was carried out by two reviewers (PH, LAK) independently, using an inductive coding approach. One author generated sub-codes and main codes, which were reviewed by the second reviewer independently. Discussions helped to resolve any disagreement around the coding scheme. A further review of the coding scheme generated descriptive and analytic themes. Then, these themes were reviewed by the second reviewer independently, with further discussion of any discrepancies.

### Sample validation

Our sample consisted of 1003 participants, 494 of whom were interviewed at both T0 and T1. We validated this sample along two dimensions. First, we showed that our sample, including the selected sample of participants who responded to both surveys, gives a useable picture of fruit and vegetable consumption in the population in the West Midlands. Second, we showed that there was a substantial change in living and working arrangements between T0 (during lockdown) and T1 (post lockdown) periods, in which participants were interviewed.

Our participants were recruited through an online survey company, and our sample is therefore not a representative sample from the population. To show that our data nevertheless paint an accurate picture of outcomes in the population, we compare demographic characteristics and fruit and vegetable consumption in our data to the Health Survey for England. This comparison is shown in table 1 below.

**Table 1:** Summary statistics.

| Sample | HSE, UK<br>(2017) |  | HSE, W Midl |  | Survey<br>(T0, 2020) |  | Survey, matched<br>(T0 and T1) |  |
| --- | --- | --- | --- | --- | --- | --- | --- | --- |
| N (nr of observations) | 9,982 |  | 982 |  | 1,003 |  | 494 |  |
| Variables | mean | stdev | mean | stdev | mean | stdev | mean | stdev |
| % female | 55 | 50 | 54 | 50 | 63 | 48 | 62 | 49 |
| Age (yrs) | 42.7 | 24.5 | 42.5 | 24.4 | 35.2 | 12.8 | 38.3 | 13.6 |
| % overweight | 32 | 46 | 33 | 47 | 27 | 44 | 30 | 46 |
| % obese | 22 | 42 | 26 | 44 | 16 | 37 | 17 | 38 |
| % sev obese | 3 | 17 | 3 | 18 | 3 | 16 | 2 | 15 |
| F&V (por/day) | 3.70 | 2.77 | 3.24 | 2.70 | 4.61 | 4.08 | 4.65 | 3.95 |
| Gender stratified statistics |  |  |  |  |  |  |  |  |
| Variables | Females | Males | Females | Males | Females | Males | Females | Males |
| Age (years) | 43.0 | 42.3 | 42.8 | 42.2 | 35.1 | 35.6 | 38.1 | 38.6 |
| % overweight | 29 | 35 | 30 | 36 | 22 | 35 | 22 | 42 |
| % obese | 22 | 22 | 27 | 25 | 17 | 14 | 19 | 15 |
| % sev obese | 4 | 2 | 4 | 2 | 4 | 1 | 4 | 0 |
| F&V (por/day) | 3.82 | 3.54 | 3.47 | 2.98 | 4.70 | 4.48 | 4.95 | 4.19 |
| Salad (por/day) | 0.43 | 0.36 | 0.37 | 0.30 | 0.47 | 0.42 | 0.50 | 0.40 |
| Pulses (por/day) | 0.26 | 0.28 | 0.25 | 0.24 | 0.50 | 0.63 | 0.52 | 0.69 |
| Veg, other (por/ day) | 0.84 | 0.81 | 0.75 | 0.66 | 1.78 | 1.68 | 1.88 | 1.42 |
| Veg, dish (por/day) | 0.13 | 0.12 | 0.14 | 0.10 | 0.30 | 0.21 | 0.26 | 0.15 |
| Fruit juice (por/ day) | 0.31 | 0.33 | 0.27 | 0.27 | 0.48 | 0.57 | 0.51 | 0.52 |
| Fresh fruit (por/ day) | 1.64 | 1.45 | 1.49 | 1.24 | 1.43 | 1.44 | 1.55 | 1.51 |
| Dried fruit (por/ day) | 0.12 | 0.11 | 0.08 | 0.09 | 0.06 | 0.07 | 0.08 | 0.07 |
| Frozen fruit (por/ day) | 0.01 | 0.01 | 0.02 | 0.00 | 0.10 | 0.07 | 0.11 | 0.05 |
| Tinned fruit (por/ day) | 0.02 | 0.03 | 0.02 | 0.02 | 0.05 | 0.05 | 0.05 | 0.04 |
| Fruit dish (por/ day) | 0.06 | 0.06 | 0.09 | 0.07 | 0.04 | 0.04 | 0.03 | 0.03 |

W Midl; West Midlands Stdev: standard deviation; F&V: fruit and vegetable; por: portion; sev: severely; Veg; vegetables

Participants in our sample are younger and more likely to be female compared to the population of the West Midlands according to the HSE. However, we match relatively well the prevalence of overweight and obesity, and patterns in fruit and vegetable consumption, both over subcategories of fruit and vegetables, and between sexes. The distribution of fruit and vegetable consumption is very similar as well, see supplement 2. There is very little evidence of sample selection in the follow-up survey with respect to the baseline survey.

### Survey timepoints

The dates of our two surveys were chosen at the end of the periods when the UK was under a national lockdown in spring 2020, and when restrictions where relatively relaxed over the summer and early autumn. The baseline survey was launched (and all responses were gathered) on 27 May, after two full months of national lockdown, which started on 23 March. This was two weeks after the government announced the first steps to lifting some of the restrictions (from 13 May more people were actively encouraged to go back to work), but before schools and businesses started to reopen (on 1 June). Responses to the follow-up survey were collected on 23 September, on the same day of the week (Wednesday), after four months of relatively relaxed restrictions and 9 days after restrictions started to be reintroduced (the “rule of 6” was introduced on 14 September).

The changes in national policy between the dates of the baseline and follow-up surveys are reflected in substantial differences in working and living arrangements reported by our participants. Among people working during both surveys, the percentage that was working from home dropped from 75 to 62% between the baseline and follow-up surveys, of which 94 versus 82% were working at home full time. The actual difference in working arrangements was even larger than these numbers suggest, because many participants lost their job between the first and second surveys. Including unemployed workers, the percentage of workers that were based at home dropped from 76 to 60% (66 vs 29% full time). The way that participants reported to do their shopping changed dramatically as well. Home delivery (including by volunteers) doubled from 14 to 29% during the lockdown, and then fell back slightly to 23% in our sample. These substantial changes in working and living arrangements make it worthwhile to explore changes in fruit and vegetable consumption and other outcomes as well.

## RESULTS

### Quantitative insights

Changes in outcomes between T0 and T1, as well as T0 versus retrospective self-report of before lockdown are presented in Figure 1. Based on the fruit and vegetable portions that participants reported consuming at T0 and T1, the average change in consumption of fruit and vegetables after the lockdown restrictions were released is not significant. Changes were computed for T1 relative to T0, so that a positive sign corresponds to an increase after lockdown ended, and thus a lower level during lockdown. For fruit and vegetable consumption, the sign is negative, indicating that consumption was similar or, if anything, higher (by a quarter portion per day) during lockdown compared with after lockdown. This is true for both total fruit and vegetable consumption, and across all its subcategories. Participants were also asked at T1 whether they had changed their consumption of fruit and vegetables since T0, and by how much. This was also insignificant, but with the change slightly positive, indicating that participants reported to have had (slightly) lower consumption during lockdown.

**Figure 1.**
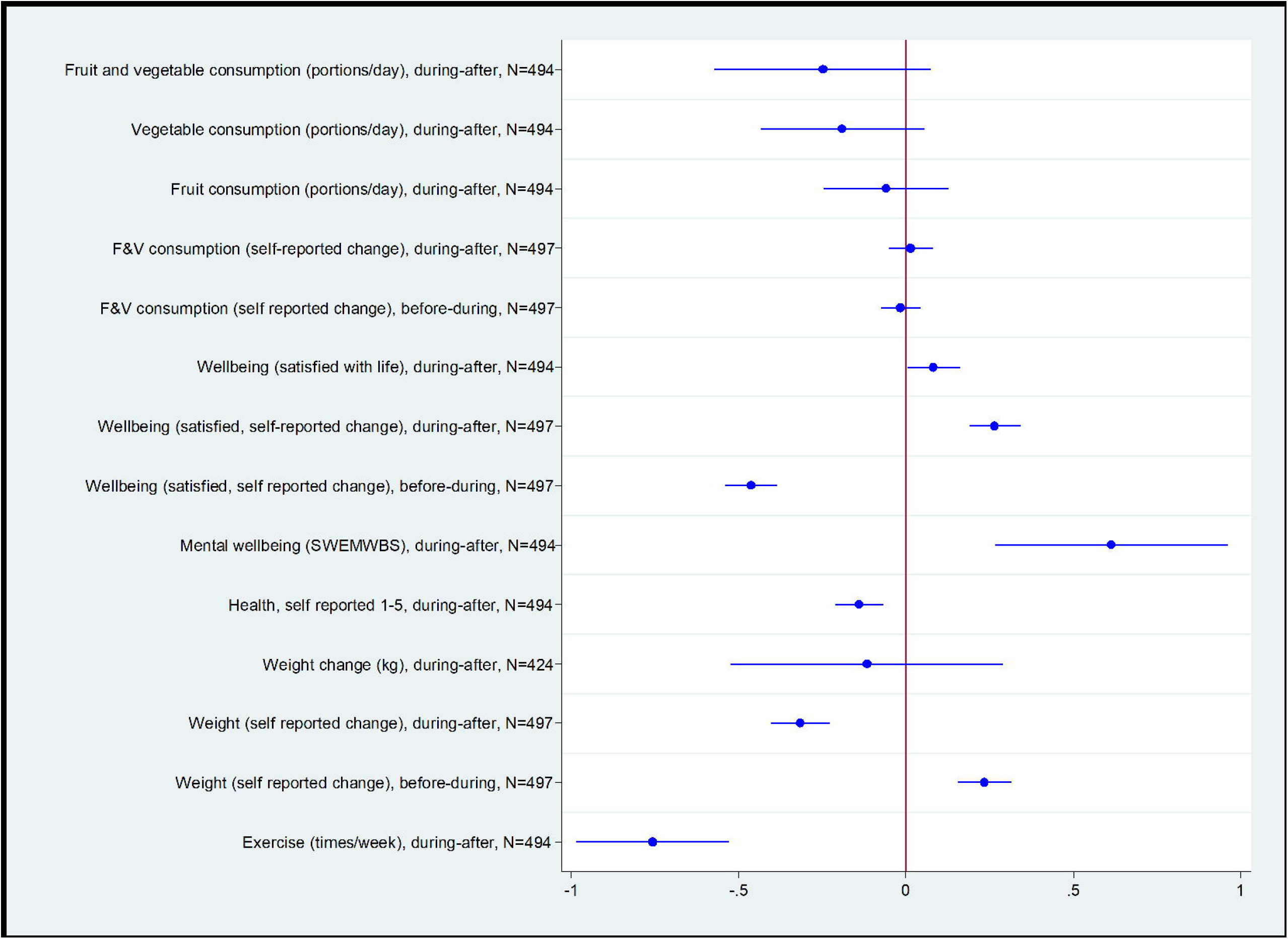
Changes in fruit and vegetable consumptions and other outcomes during lockdown.

Figure shows point estimates and 95% confidence intervals

We examined whether the change in fruit and vegetable consumption between T0 and T1 different in specific population subgroups, with the findings presented in Figure 2.

**Figure 2.**
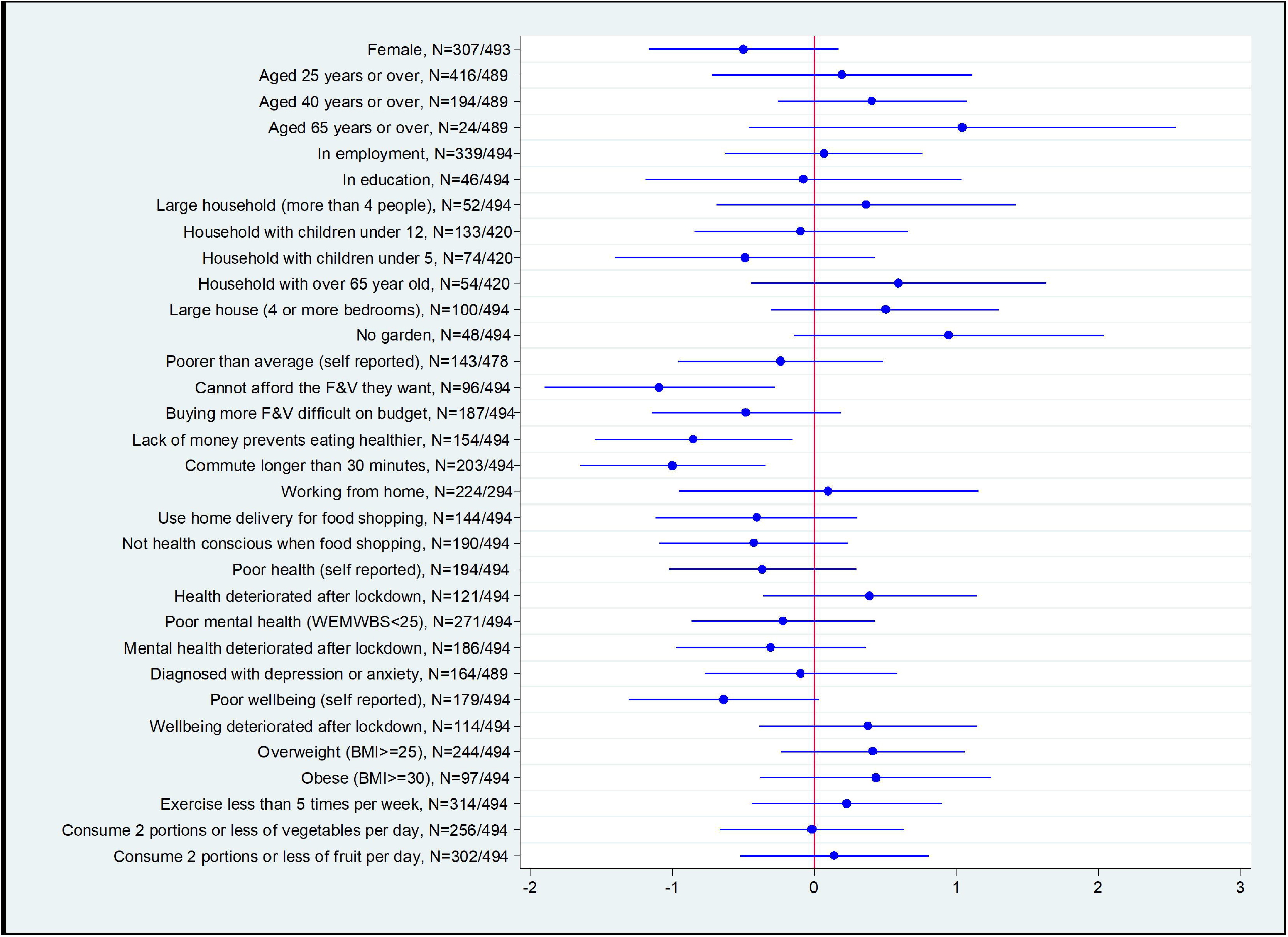
Heterogeneity in change (after versus during lockdown) in fruit and vegetable consumption (portions/day)

Figure shows point estimates and 95% confidence intervals

Most of the variables that we consider do not seem to be associated with differences in the change in fruit and vegetable consumption. The only characteristics for which we observe different changes in consumption during lockdown that are significant at the 5% level are some of the budget measures (“cannot afford to buy the fruit and vegetables I want in my usual shops” and “lack of money prevents me from eating healthily”) and having a long commute (more than 30 minutes), which associate with a larger drop in consumption after lockdown ended, indicating a larger increase in consumption during lockdown. We also found that female participants and participants reporting low levels of wellbeing may have experienced higher increases in fruit and vegetable consumption during lockdown, and older people (and perhaps participants with overweight or obesity as well) may have increased their consumption less, or even consumed less fruits and vegetables, during lockdown. These differences across gender and budget are not statistically significant, but the effect sizes are large, and neither men nor participants who did not feel their budget was tight reported any difference in their fruit and vegetable consumption.

Based on the descriptive evidence as reported above, we identified characteristics that may be associated with higher or lower fruit and vegetable consumption during lockdown and explored these further in a multivariable analysis. In particular, we regressed the change in fruit and vegetable consumption on measures of a low budget for food shopping, age and overweight as potential risk factors, and we estimate the regression separately for women and men. The results are reported in Table 2.

**Table 2:**
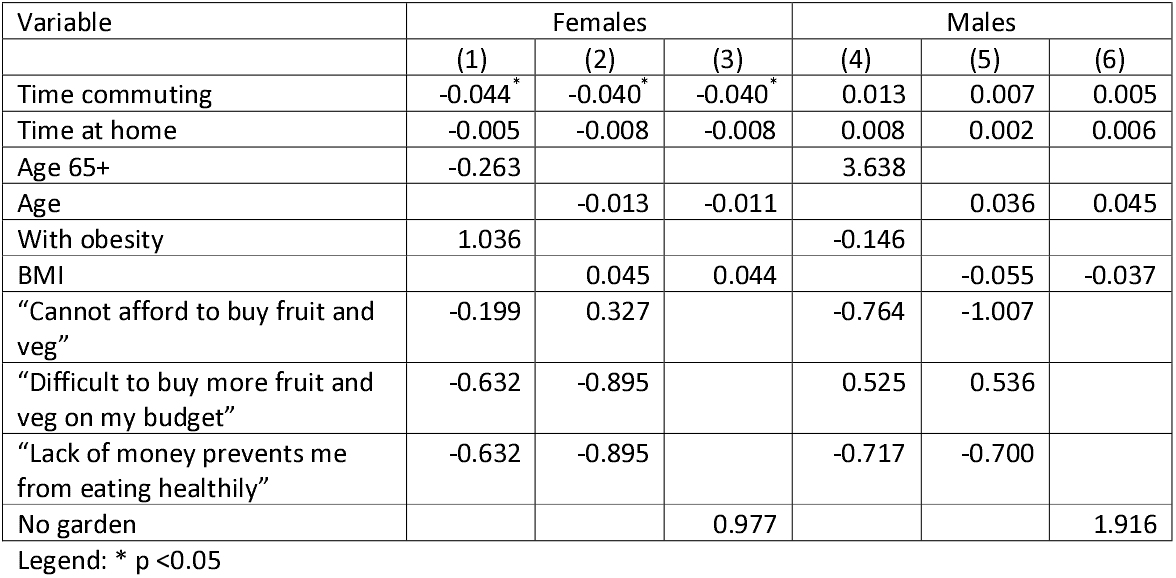
Multivariable analysis.

| Variable | Females |  |  | Males |  |  |
| --- | --- | --- | --- | --- | --- | --- |
|  | (1) | (2) | (3) | (4) | (5) | (6) |
| Time commuting | -0.044* | -0.040* | -0.040* | 0.013 | 0.007 | 0.005 |
| Time at home | -0.005 | -0.008 | -0.008 | 0.008 | 0.002 | 0.006 |
| Age 65+ | -0.263 |  |  | 3.638 |  |  |
| Age |  | -0.013 | -0.011 |  | 0.036 | 0.045 |
| With obesity | 1.036 |  |  | -0.146 |  |  |
| BMI |  | 0.045 | 0.044 |  | -0.055 | -0.037 |
| “Cannot afford to buy fruit and veg” | -0.199 | 0.327 |  | -0.764 | -1.007 |  |
| “Difficult to buy more fruit and veg on my budget” | -0.632 | -0.895 |  | 0.525 | 0.536 |  |
| “Lack of money prevents me from eating healthily” | -0.632 | -0.895 |  | -0.717 | -0.700 |  |
| No garden |  |  | 0.977 |  |  | 1.916 |

Legend: \* p < 0.05

In order to capture the difference between participants with a long versus no or a short commute, we hypothesised that this may be due to participants using the time saved commuting during lockdown to cook. To test this hypothesis, we include a measure for time saved in the regressions. Time saved commuting is calculated by multiplying a respondent’s usual commute in minutes with the change in the fraction of time working from home.

The first three columns in table 2 refer to female, the last three columns refer to male participants. The other differences across columns are in the measures we entered into the models to examine the effect of age and overweight (dummies for over 65 years and obese versus age and BMI and tight budget (dummies for “cannot afford to buy the fruit and vegetables I want in my usual shops”, buying more fruit and vegetables would be difficult on my budget” and “lack of money prevents me from eating healthily” versus dummy for “no access to a garden or outdoor space”).

The multivariable results suggest that it was mostly female participants who saved time commuting who increased their consumption of fruit and vegetables during lockdown. Adjusting for time saved commuting, time spent at home is not a significant determinant of fruit and vegetable consumption. Neither time saved commuting nor time spent at home matters for fruit and vegetable consumption reported by male participants.

Other than the effect of time saved commuting, we find no evidence for any of the potential determinants of diets that we explored, except perhaps that older men may have had lower consumption of fruit and vegetables during lockdown. There does not seem to be any association at all for obesity and overweight, nor for measures of low income or tight food budgets, for either women or men.

### Wellbeing outcomes

As we may have expected, subjective wellbeing and mental wellbeing was lower during lockdown than afterwards [Figure 1].

In the first survey during lockdown, participants rated their life satisfaction on average lower by almost 0.1 point, and this change was significant (p = 0.039). The same trend emerges from a retrospective question from the second survey after lockdown was released, asking participants to compare their lives during and after to before the lockdown, again on a 5-point scale, this time ranging from “much less satisfied” (1) to “much more satisfied” (5). In hindsight, participants rated their life satisfaction almost 0.5 points lower (on a 1-to-5 scale) during the lockdown, which partially, but not fully recovered after lockdown was eased. Both of these changes are statistically significant (p-value <0.01). Lower wellbeing during lockdown manifested itself as lower mental wellbeing measured using SWEMWBS as well. After lockdown ended, the reported mental wellbeing score was on average 0.6 points higher than during lockdown, and this change was statistically significant (p-value <0.01). Young people in full-time education suffered the largest drop in wellbeing during the lockdown, see Figure 4 in supplement 3.

### Health and weight

Lower wellbeing, however, did not translate to worse self-reported health overall, nor to self-reported weight gain. Health, self-reported on a scale from “very bad” (1) to “very good” (5), was higher during lockdown, based on the significant decrease by 0.14 points when lockdown restrictions were released. Self-reported weight also decreased, by 0.1 kg on average, when lockdown ended. Although this change is not significant, it is consistent with participants retrospectively reporting to have lost weight during lockdown and gained it again when lockdown ended, (retrospective weight change is shown in Figure 1. This was reported by participants on a 5-point scale ranging from “gained a lot” (1) to “lost a lot of weight” (5), so that positive changes indicate weight loss). Physical activity also decreased significantly (p-value <0.01) after lockdown ended, indicating participants on average exercised 0.8 times per week more often during lockdown. Combined with our earlier finding that fruit and vegetable consumption increased most for women who saved the most time commuting, this result suggests that healthy behaviours may have improved during lockdown because people had more time.

## Qualitative insights

The free text questions were completed by 330 participants. Qualitative data covered two main topics, change in fruit and vegetable consumption and weight change.

### Fruit and vegetable consumption during lockdown

Participants reported various reasons for reduced fruit and vegetable consumption during lockdown, with two main emerging themes: access issues and behavioural change. Other participants reported attitudes with two emerging themes: abundance of resources and successful change. These both associated with increased fruit and vegetable consumptions.

### Access issues

Participants reported various factors that affected their usual shopping habits and access to shops or food stores. Change in the mode of shopping was a barrier to fruits and vegetable purchases. For instance, online delivery slots were not as frequent as “usual” shopping behaviour, the pickers may not have chosen fruit and vegetables according to shelf life as participants would have done if they were choosing the products themselves in the shop, and the long waiting time to receive a food delivery had a knock-on effect on shelf life.

> *“Delivery meant food didn’t last as long as I couldn’t select it. Also, I was often stuck with one delivery every 2-3 weeks and fresh fruit and veg tends not to last that long”*

Participants reported some of the challenges such as infrequent shop visits and the short shelf life of fruits and vegetables, consequently people ran out of items before their next food shop

> *“Shopping for food was done less regularly and fresh fruit and vegetables did not last the whole time between shopping trips”*
>
> *“I think this is down to not going out to the shops as frequently, I’ve tended to buy only once or twice a week, whereas before covid 19 I used to shop almost daily for fresh salad and veg and fruit”*

Although there were alternatives such as frozen and canned items, participants generally preferred fresh produce.

> *“Access to fresh fruit and veg was restricted as was availability in shops. Not as keen on frozen and dislike tinned”*

People were not comfortable with online selections as they did not have the choice to inspect items or received items of poor quality.

> *“I was ordering online from a supermarket and the fruit and vegetables picked by them for delivery were of poor quality and put me off eating fruit”*

There was a frequent reporting of financial barriers to purchasing fresh produce. They also raised concerns around the costs of fresh fruits and vegetables, for instance one participant could not justify the costs of items with a short shelf life.

> *“Shops are more expensive and fresh food goes off too quick so I am unlikely to buy unless I want it that day”*.

Others did not have the financial means for regular purchases of fresh produce

> *“Ordering online with minimum spend made it hard to top-up fresh veg and especially fresh fruit”*.

Worries were reported around the risk of infection from purchasing fresh produce, one participant reported

> *“Less money and more afraid of the fruit market so haven’t bought from commercial stores”*.

Concerns around income and job insecurity was another barrier

> *“I had no work during lockdown and wasn’t furloughed, so money was an issue”*.

### Negative behavioural change

The behaviour of participants changed during national lockdown, with possible contributors including lack of motivation, increased stress, deterioration in mental health and lack of general interest. There was a consistent migration towards unhealthy dietary habits. Regarding emotional eating, associated factors included boredom, feeling low, stress and economic uncertainty.

> *“during lockdown I put on a lot of weight as I was eating a lot as I was bored”*
>
> *“The lockdown period and the current period have been very stressful due to the economic impact on businesses and this has often meant turning to unhealthy comfort food”*

People found comfort in takeaways, calorie dense food and snacking

> *“food was one of the only things that could uplift my mood in lock down so I ate a lot more junk food and didn’t think about how much fruit and veg I ate. However I did go through a phase of making healthy wholesome meals. But I still snack badly”*

Some lost the motivation “can’t be bothered” to dedicate efforts around appearance, cooking, preparing fresh meals or following a healthy lifestyle.

> *“I can’t be bothered to cook anymore because I care less about my appearance now I don’t have to go anywhere”*
>
> *“During lockdown and now we have been feeling lazy and seemed to prefer ready cooked meals and takeaways rather than preparing fresh food”*
>
> *“was doing Slimming World before lockdown and following a healthy diet, but not so much at the moment”*

Change in working arrangements negatively affected participants’ food choices. Working from home changed eating habits. Some participants reported adding fruit to their packed lunch pre-lockdown, with fewer snacks and more dietary structure:

> *“When I go to work I take my dinner along with fruit, I always eat the fruit that I take to work with me but when I am at home and there are alternative snacks I generally don’t seem to pick fruit even though I do try to maintain a reasonable fruit intake”*
>
> *“wasn’t following my normal routine every day so habits changed”*

### Abundance of resources

A number of participants highlighted several resources that helped them increase their fruit and vegetable consumptions. These included social support, better access, improved knowledge and more time. Social support involved a reliance on close members to help with the food shop and having family members to help with the cooking.

> *“my sister was doing my shopping and she brought me more, the children were home all the time and they love home-made soups and smoothies, as well as fresh fruit and Greek yoghurt, without the school run and after school clubs I had more time to cook properly”*

Others highlighted the positive influence of living with others as they followed healthier dietary habits.

> *“My housemate has taken on the shopping and cooking. He eats vegan and a strict “clean eating” diet so he likes to cook something for all of us to share most days. As I don’t want to go out to supermarkets, I am basically following his diet, with maybe a bit of extra bread. The meals are mostly stews, curries etc based on pulses and veg”*.

Having additional support with childcare seemed to improve eating habits

> *“My partner was around to help out with the children. Now he’s back at work I easily forget to snack on fruit and eat meals if my days are busy”*

Regular access to healthier options such as fruit and vegetable boxes, local farm shops and growing fresh produce helped people to increase their consumption.

Arranging for regular supplies such as deliveries provided more options to eat and cook from

> *“got fruit and veg box delivery during lockdown so had more veg in the house to choose to cook”*

while others grew their own produce

> *“We started to grow our own vegetables. Plus we had apples in the garden. So this upped our veg consumption”*

The change in mode of shopping where some chose to walk to shops or access their local market positively influenced participants’ purchases

> *“During lockdown we were walking to local farm shop and buying more fruit. During lockdown we walked to get shopping so bought as much healthy food we could carry. Now we drive to supermarkets so can get whatever we want”*.

Many participants report to have had more time to organise their meals, increase their health awareness and prioritise their wellbeing. Participants had more time to consider what they purchase online or more time dedicated to the thinking process of meal preparations.

> *“I was on furlough and did more cooking, a lot of which were vegetarian dishes, I also purchased a soup maker and I made a lot of vegetable soups with it”*
>
> *“More time and energy to put an effort into eating well and looking after my health and wellbeing, which can sometimes otherwise be lower down in my day-to-day list of priorities”*

### Successful change

Many participants reported a clear and strong motivation to become healthier, acquire better habits and lifestyle. Some wanted to build a healthier and stronger immune system in the light of the Covid-19 pandemic

> *“I think I was more conscious of being healthy and wanting to boost my immune system”*
>
> *“More motivated to eat healthy to try and combat COVID-19”*

while others wanted to protect their mental health

> *“I got fed up of eating junk food and try to eat healthier to improve my mental health”*.

The national lockdown was a time of reflection to some and a “reality check” to others. Participants reported a conscious effort to improve dietary intakes and reduce weight.

> *“I had a reality check when I stepped on the scales and realised I needed to improve my diet. My nutribullet was the best lockdown purchase I made. I now start my day with a breakfast smoothie rather than chocolate and my weight is going down and I feel much better”*

Although some food outlets were closed but this seemed to have a positive effect on participants’ habits

> *“most takeaways were shit during lockdown so only option was to cook”*

The challenges in “usual” shopping modes seemed to serve as enablers to healthier shopping habits

> *“during lockdown we had to plan our shopping trips more thoroughly and therefore it was easier to buy more veggies and fruit. we were influenced by the trend of getting more healthy so we ate better”*.

### Fruit and vegetable consumption after lockdown

Two emerging themes were associated with decreased fruit and vegetable consumption: lockdown behavioural spill over and external and internal distress. Increased fruit and vegetable consumptions was associated with having improved access and increased health consciousness.

#### Lockdown behavioural spill over

Some of the contributes of behavioural change during national lockdown such as lack of motivation and deportation in mental health seemed to spill into the post lockdown period.

> *“My motivation has dropped since earlier in the year, lockdown has caused a lot of mental health issues to me personally”*

Lockdown behaviours such as infrequent shopping trips and working from home arrangements negatively affected participants’ dietary consumptions

> *“Before lockdown, when I worked in the office, I would make sure I had a healthy lunch, so would go to subway for a salad or pick something up at a supermarket. Now that I work from home, I tend to leave the house less and so make do with whatever food is in the cupboards. We don’t tend to buy a lot of fresh produce at the moment so that it doesn’t go to waste”*

Other found it difficult to release themselves from lockdown habits despite attempts to become healthier

> *“Started to go back to the gym but it is hard breaking the lockdown habits”*

while some questioned the back to normal climate

> *“We have not yet returned to a pre-lockdown state”*.

#### External and internal distress

Lack of financial means, time constraints and access issues were external factors associated with reduced fruit and vegetable consumptions. Participants reported financial barriers and inability to regularly afford fruit and vegetables. In addition, participants found it difficult to frequently access shops for fresh produce

> *“I don’t like having to go shopping wearing a mask so am less likely to nip into a supermarket for a top up shop when I’ve run out of fruit. I used to buy more expensive types of fruit when they were reduced at the end of the day and it is now harder to do that as supermarkets are offering less reductions. And working from home makes me want to snack on less healthy foods when I get bored”*

Comfort eating, poor mental health and lack of motivation were internal factors that were associated with reduced consumptions

> *“More treats and snacks now to try and get a boost. Wrong thing to do, but difficult to resist”*

#### Improved access

Participants reported changes in shopping habits, such as more frequent visits to the supermarkets and buying more fresh produce.

> *“I am comfortable visiting supermarkets more often, so I buy more fresh produce”*

Others reported that the continued use of online shopping or deliveries helped them to plan and eat healthier.

> *“I have continued to order food online, and so have been able to continue the eating habits started during lockdown. Also, I am still working from home, so have the opportunity to plan and cook meals in a better way”*

Improved access to facilities for exercise, both indoors and outdoors, helped to increase activity which in turn drove desire to be healthier.

> *“Gone back to gym and PT sessions so eating healthier”*
>
> *“Since lockdown rules had eased, I was able to do more of my daily exercises outdoors which at the same time made me want to eat more healthily”*

Some participants had increased access as a result of home-grown vegetables and fruit during summer.

> *“We have the time to go foraging for blackberries and have grown our own fruit and vegetables”*

### Increased health consciousness

Participants became more conscious of their health and wanted to increase fruit and veg consumption in order to improve immunity, lose weight or start leading healthier lifestyle.

> *“Trying to boost the immunity that in case covid-19 happens to me then at least i can go down trying to fight with it. I mean i want to give myself a fighting chance”*
>
> *“I have a motivation to eat healthily as I have put on weight since lock down and I want to lose it before starting university”*
>
> *“Motivation to finally take better care of my health and change my habits”*

### Weight changes

Participants were asked to describe changes to their weight during and after the national lockdown. Participants were divided into three main categories (overall weight gain, weight control and weight loss followed by weight gain) based on changes to their weight during the two time points.

#### Weight loss followed by weight gain

Eight participants experienced weight loss during lockdown and weight gain after lockdown finished. This was as a result of having less time to exercise after lockdown finished as well as having less time to look after health when period of lockdown finished.

> *“I had a lot more time to exercise/focus on my body during lockdown so lost weight and gained muscle”*
>
> *“I am busier after lockdown and am paying less attention to diet”*

#### Overall weight gain

137 participants experienced some degree of weight gain, either during lockdown, after lockdown finished or during both. The most common theme was increase in alcohol, snacking and unhealthy food, secondary to mental health problems or boredom.

> *“Depression, anxiety due to lockdown. Now total lack of motivation, no money, no exercise. Vicious cycle of depression and comfort eating then weight gain filled by guilt and more comfort eating. I know what I should eat but I can’t afford to do it. Then fat shammed by society so can’t see the point of trying anymore”*
>
> *“More time without purpose and boredom”*
>
> *“I have baked a lot more and made cakes! I have also increased the amount of alcohol I drink and look forward to “ opening time” for a glass of wine I have watched more television and therefore been sitting more frequently I have not been able to swim my 30 lengths twice a week”*
>
> *“Struggle with emotions which causes me to snack and rubbish food”*

Some reported a change in health status, such as new medical condition or pregnancy.

> *“I was pregnant during lockdown. I am still carrying baby weight now. Due to have two small children I also skip means and sometimes snack on junk food”*

Some experienced overall reduction in activity.

> *“Both me and my husband were juggling working from home and childcare. If usually do exercise on my way fhe from work/ before picking kids up but felt time was always being used with work/ childcare/ housework and not much left in between”*
>
> *“Less activity than normal due to shielding and boredom eating”*

#### Weight control

185 participants described overall weight loss during lockdown or post lockdown. The facilitators for the positive changes were improved diet with portion control and overall healthier foods.

> *“Eating healthier as I was home cooking more or less all my food consumption. So I was able to manage all the ingredients that went into preparing my food. Also I tried to reduce carb intake, which proved very helpful in weight loss”*

Participants described increase in motivation after lockdown ended.

> *“I was trying to lose weight before lockdown, then comfort ate at the beginning of lockdown but then decided to get back on track especially as being overweight can hinder you if you catch covid 19”*
>
> *More exercise to combat stress, not working doing my physical job so not needing to eat as much. Having more sleep and more energy to do the fitness I wanted*.

Increase in physical activity also played a role.

> *“I have been cycling to work and back since i went back after lock down to avoid public transport”*
>
> *“I challenged myself to focus on my running and ran my first half marathon during lockdown on the treadmill”*

Change in eating habits with less take away and eating in restaurants.

> *“I believe less meals out and nights out drinking has also helped me lose weight”*

Stress induced loss of appetite was one of the reasons for weight loss.

> *“Partly anxiety causing appetite loss and also cutting right down on drinking and snacking less”*

## DISCUSSION

Our study found that fruit and vegetable consumption of residents of the West Midlands during lockdown was not significantly lower than outside of lockdown. In fact, our evidence suggests consumption may have been slightly higher during lockdown based on contemporary reports of portions consumed during the lockdown (T0: May) and after the lockdown (T1: September). We also asked at T1 whether people had increased or reduced their fruit and vegetable consumption since T0 and the answer was a non-significant decrease.

We hypothesised that lockdown would have negative effect on diet including fruit and vegetable consumption, which has been evidenced in other published articles many of which have used cross-sectional designs (15, 30). When asked to self-report changes, our study participants expressed the opinion that their fruit and vegetable consumption had declined (quantitatively non-significantly, but explicity in the qualitative data). However, we did not find support for the hypothesis in our contemporaneous quantitative data on portions consumed at two time points. Another longitudinal study using five British cohort studies has similar findings to our study, in that fruit and vegetable intake was broadly similar pre and during lockdown (31). This raises implications for design and interpretation of studies on this topic, suggesting that studies asking participants to self-report increases or decreases in fruit and vegetable consumption may give inaccurate results.

We explored whether there were particular population subgroups whose fruit and vegetable consumption were more adversely or less adversely effected by lockdown. We found that (i) some measures of financial constraints and (ii) having a long commute (outside of lockdown) are associated with higher consumption of fruits and vegetables during lockdown, than afterwards. There is weaker evidence that women and participants reporting low levels of wellbeing may have experienced higher increases in fruit and vegetable consumption during lockdown, whereas older people may have experience lower consumption during lockdown than afterwards. A multivariable analysis revealed that the patterns for women can largely be explained by time saved commuting during lockdown (compared with afterwards), while no other variable investigated was significantly associated with change in fruit and vegetable consumption. For men, being older was (weakly) associated with lower consumption of fruit and vegetables during lockdown than afterwards, but no other variables were significantly associated.

A previous high-quality longitudinal study of the UK population, using five British birth cohorts, found that younger cohorts were more likely to increase fruit and vegetable consumption during lockdown than older cohorts, in line with our findings (31). We believe we are the first UK study to report a differential effect based on time saved commuting, which may be highly policy relevant, not just in the context of ‘lockdowns’ but for increasing fruit and vegetable consumption in the population generally. Previous studies have reported particularly adverse effects of lockdown on diet for populations with overweight and obesity that we did not identify (30, 32). The difference between their findings and ours may be due to the fact that these previous studies were mainly cross-sectional.

In terms of our secondary outcomes, firstly, wellbeing was lower in lockdown, measured by either life satisfaction or SWEMWBS. The changes during lockdown are also very large and broadly greater in size than the unhappiness associated with being unemployed or maritally separated, which are well-documented as the two strongest depressants of reported happiness in ‘Western’ society (33, 34). The reported drop in overall life satisfaction, is also in line with previous studies, for example a longitudinal analysis of the UK Household Longitudinal Study found that psychological distress increased one month into lockdown from 19.4% in 2017-2019 to 30.6% in April 2020 (35) and the percentage of UK adults experiencing a significant mental health problem is estimated to have risen by approximately 50% based on nationally representative data collected before and during COVID-19 lockdown (36). An additional study showed that aggregate wellbeing in the UK fell by 0.65 points on the same scale (37). In contrast, we found that self-reported health was higher in lockdown than after lockdown which may have be driven by the trend towards increased fruit and vegetable consumption and statistically significant increase in physical activity during lockdown compared with after lockdown. Lastly, self-reported weight was higher during lockdown than after lockdown but this was non-significant.

Finally, we explored some of the barriers and facilitators to fruit and vegetable consumption during lockdown and found that access issues, and negative behaviour change reduced fruit and vegetable consumption while abundance of resources and successful behaviour change which were associated with increased fruit and vegetable consumption

The major strength of this study is that we have longitudinal data from the same participants allowing more objective analysis of dietary change than studies asking about retrospective habits at one point in time. Few studies have been published with a longitudinal design. A further strength of our study is the qualitative data collection and analysis. We could not identify any other qualitative or mixed methods study that has been published in the literature on this topic from anywhere in the world. This study additionally has limitations. We recruited participants online, which means the sample is not representative of the population of the West Midlands, although as described in our methods our sample appears to be quite similar to a representative sample (the HSE) in terms of socio-demographic characteristics and patterns of fruit and vegetable consumption. Nevertheless, it is possible that selection bias affected our results. For example, people whose time was severely negatively affected by the lockdown, e.g. essential workers with increased burden of work or parents with small children staying home from school, may have been less likely to respond to an online survey and may therefore be underrepresented in our data.

## CONCLUSIONS

There seems to be no evidence for a negative effect of lockdown on physical health, in terms of behavioural risk-factor for disease (fruit and vegetable consumption, physical activity-both of which were maintained or increased during lockdown for our study sample) and in terms of self-reported weight and self-rated health. On the other hand, there is clear evidence that lockdown was detrimental to mental health.

We did not find evidence of significant inequalities in the effect of lockdown on fruit and vegetable consumption between population subgroups explored-although there is some indication that women who usually have a long commute were particularly able to increase their fruit and vegetable consumption during lockdown and perhaps that older men were more vulnerable to reduced fruit and vegetable consumption during lockdown.

Concerns that lockdown negatively affected diet quality in the UK may be allayed to some extent by our findings, however, an adverse effect on mental wellbeing is clearly apparent from our data.

## Supporting information

Supplement 1A. Questionnaire survey T0

Supplement 1B. Questionnaire survey T1

Supplement 2. Distribution of fruit and vegetable consumption

Supplement 3. Heterogeneity in menal wellbeing

## Data Availability

Data available from the corresponding author upon reasonable request and subject to the conditions of the ethical approval.

## LIST OF ABBREVIATIONS

BMI: Body Mass Index
F&V: Fruit and vegetable
HSE: Health Survey for England
ONS: Office for National Statistics
Por: Portion
Sev: Severely
Stdev: Standard Deviation
SWEMWBS: Short Warwick-Edinburgh Mental Wellbeing scale
UK: United Kingdom
Veg: Vegetables
W Midl: West Midlands

## DECLARATIONS

### Ethics approval and consent to participate

We received ethical approval under the DR@W2 agreement from the Humanities and Social Sciences Research Ethics Committee (HSSREC) at the University of Warwick (reference number 168/19-20).

### Consent for publication

Not applicable

### Availability of data and materials

The datasets used and/or analysed during the current study are available from the corresponding author on reasonable request.

## Authors’ contributions

The idea for this study was conceived in a group meeting with all authors. TvR designed the survey with input from the other authors. TvR did the quantitative analysis of the data with help from LW. LAK and PH did the qualitative analysis. OO drafted the manuscript with input from the other authors. All authors read and approved the final version.

## Acknowledgements

The authors would like to thank Maartje Kletter for excellent research assistance.

