## Supplement 1A. Questionnaire survey T0 for "Healthy diets, lifestyle changes and wellbeing during and after lockdown: Longitudinal evidence from the West Midlands"

Eating behaviour during Covid lockdown - Prolific

Survey Flow

EmbeddedData

PROLIFIC_PIDValue will be set from Panel or URL.

Standard: Participant Information Sheet (2 Questions)

Branch: New Branch

If

If PARTICIPANT INFORMATION SHEET   Study Title: The impact of lockdown on health and wellbeing and f... I do not wish to participate Is Selected

EndSurvey: Advanced

Branch: New Branch

If

If PARTICIPANT INFORMATION SHEET   Study Title: The impact of lockdown on health and wellbeing and f... I have read the above and consent to take part in this study Is Selected

Block: Lockdown situation. (12 Questions)

Standard: Physical and mental health (6 Questions)

Standard: Fruit and vegetable consumption (23 Questions)

Standard: Demographics (11 Questions)

EndSurvey: Advanced

EndSurvey:

| Page Break |
| --- |

Start of Block: Participant Information Sheet

Q53
**PARTICIPANT INFORMATION SHEET**
 
**Study Title:** The impact of lockdown on health and wellbeing and fruit and vegetable consumption
 
**Investigators:**
Thijs van Rens (Department of Economics)
Lola Oyebode and Lena AlKhudairy (WMS Population Evidence and Technologies)
Thomas Barber and Petra Hanson (Human Metabolism Research Unit and University Hospitals Coventry and Warwickshire)
Ioannis Nezis (School for Life Sciences, Biomedical Science)
Lukasz Walasek (Department of Psychology)
Redzo Mujcic (Warwick Business School)  
 
In this survey, we will ask you some questions about your situation during the Covid-19 lockdown, about your well-being and your mental and physical health, and about your fruit and vegetable consumption both during the lockdown and in normal times. We are interested in finding out whether there is a change in eating behaviour during the lockdown that may contribute to weight gain. We will also ask you some general questions about yourself (e.g. your gender and your age), which will help us understand if different people are affected differently.
 
This research is funded by the University of Warwick Global Research Priority on Food.
 
Your participation is completely voluntary. You can withdraw at any time, and for any reason, simply by closing your browser.  
 
No identifiable data will be collected from you as part of this study. This means that once your responses have been submitted to the research team, it will not be possible to withdraw this data as your individual responses cannot be identified. Data will be securely stored on University of Warwick computers and will be processed only for the purpose of scientific analysis. Access to the data will be restricted to the researchers. Summaries may be presented at conferences and included in scientific publications. Data will be reviewed after a period of 10 years, in line with the University of Warwick data retention policy.
 
Please refer to the University of Warwick Research Privacy Notice which is available here:
https://warwick.ac.uk/services/idc/dataprotection/privacynotices/researchprivacynotice
or by contacting the Information and Data Compliance Team at. 
 
This study has been reviewed and given favourable opinion by the University of Warwick’s Humanities and Social Science Research Ethics Committee (HSSREC).
 
If you require further information, please contact.
 
**Who should I contact if I wish to make a complaint?**
 
Any complaint should be addressed to the person below, who is a senior University of Warwick official entirely independent of this study:
 
**Jane Prewett (Head of Research Governance)**
Research & Impact Services
University House
University of Warwick
Coventry
CV4 8UW

 
If you wish to raise a complaint on how we have handled your personal data, you can contact our Data Protection Officer, Anjeli Bajaj, Information and Data Director who will investigate the matter:.
 
If you are not satisfied with our response or believe we are processing your personal data in a way that is not lawful you can complain to the Information Commissioner’s Office (ICO).
 
**If you are concerned about your mental health or wellbeing**, [these helplines and support groups](https://www.nhs.uk/conditions/stress-anxiety-depression/mental-health-helplines/) can offer expert advice.
 
**Thank you for taking the time to read this Participant Information Leaflet.**

- I have read the above and consent to take part in this study (1)
- I do not wish to participate (4)

| 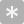 | 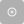 |
| --- | --- |

Q54 Please enter your Prolific ID:

________________________________________________________________

End of Block: Participant Information Sheet

Start of Block: Lockdown situation.

Q14 First, we would like to ask you some questions about the situation you are currently in. Here, we refer to the last two weeks, when we were in “lockdown” to control the Coronavirus.

Q1 What was your work/school situation prior to the lockdown?

- Going to school or college full-time (including on vacation) (1)
- In paid employment or self-employed (or temporarily away) (2)
- On a Government scheme for employment training (3)
- Doing unpaid work for a business that you own, or that a relative owns (4)
- Waiting to take up paid work already obtained (5)
- Looking for paid work or a Government training scheme (6)
- Intending to look for work but prevented by temporary sickness or injury (7)
- Permanently unable to work because of long-term sickness or disability (8)
- Retired from paid work (9)
- Looking after home or family (10)
- Doing something else (11)

Display This Question:

If What was your work/school situation prior to the lockdown? = Doing something else

Q15 Please specify your work/school situation prior to the lockdown

________________________________________________________________

Display This Question:

If What was your work/school situation prior to the lockdown? = Going to school or college full-time (including on vacation)

Or What was your work/school situation prior to the lockdown? = In paid employment or self-employed (or temporarily away)

Or What was your work/school situation prior to the lockdown? = On a Government scheme for employment training

Or What was your work/school situation prior to the lockdown? = Doing unpaid work for a business that you own, or that a relative owns

Q2 Are you currently

- Not working/studying (1)
- Working/studying from home (2)
- Working at your workplace as an essential worker (3)
- Working/studying at your workplace/school for other reason (4)
- Working/studying partly from home, partly at workplace (5)

Display This Question:

If Are you currently = Working/studying at your workplace/school for other reason

Q16 What is the reason that you are working/studying at your worksplace/school?

________________________________________________________________

Display This Question:

If Are you currently = Working/studying partly from home, partly at workplace

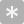

Q17 Roughly what percentage of the time are you working from home? (%)

________________________________________________________________

Q3 How many other people are there in your household, including yourself? Your household includes everyone living with you in the same house, sharing a kitchen.

- Children 0-4 years (1) ________________________________________________
- Children 5-11 years (2) ________________________________________________
- Children 12-17 years (3) ________________________________________________
- Adults 18-64 years (12) ________________________________________________
- Adults 65-74 (13) ________________________________________________
- Adults 75+ (14) ________________________________________________

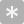

Q4 How many bedrooms does your household have?

________________________________________________________________

Q59 Do you have access to a garden or other private outdoor area?

- Yes (23)
- No (24)

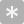

Q5 How many times per week do you exercise or go outdoors for active recreation (walk, sports, ...)?

________________________________________________________________

Q6 How do you usually travel to and from work or school?

- Walk (1)
- Run (2)
- Bicycle (3)
- Private car or motorbike (4)
- Public transportation (5)

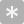

Q7 Prior to the lockdown, how long was your usual commute time per day (in minutes, both to and from work or school)?

________________________________________________________________

End of Block: Lockdown situation.

Start of Block: Physical and mental health

Q21 We would now like to ask you some questions about your health during the last 2 weeks, while we were in lockdown to control the Coronavirus.

Q9 How was your health in general during the last 2 weeks?

|  | Very bad (1) | Bad (2) | Fair (3) | Good (4) | Very good (5) |
| --- | --- | --- | --- | --- | --- |
| (6) |  |  |  |  |  |

Q10 For each of the following four questions, please give an answer on a scale from 0 to 10, where 0 is "not at all" and 10 is "completely".

|  | 0 (not at all) (1) | 2.5 (6) | 5 (7) | 7.5 (9) | 10 (completely) (5) |
| --- | --- | --- | --- | --- | --- |
| All things considered, how satisfied were you with your life over the last 2 weeks? (1) |  |  |  |  |  |
| Overall, to what extent do you feel that the things that you do in your life are worthwhile? (6) |  |  |  |  |  |
| Overall, how happy did you feel yesterday? (7) |  |  |  |  |  |
| On a scale where 0 is "not at all anxious" and 5 is "completely anxious", overall, how anxious did you feel yesterday? (8) |  |  |  |  |  |

Q11 Compared with how satisfied you usually are with your life (when not in lockdown), would you say that over the last 2 weeks you felt:

|  | Much less satisified than usual (1) | Less satisfied than usual (2) | About the same as usual (3) | More satisfied than usual (4) | Much more satisfied than usual (5) |
| --- | --- | --- | --- | --- | --- |
| (1) |  |  |  |  |  |

Q12 Which of these best describes your experience over the last 2 weeks?

|  | None of the time (1) | Rarely (2) | Some of the time (3) | Often (4) | All of the time (5) |
| --- | --- | --- | --- | --- | --- |
| I’ve been feeling optimistic about the future (1) |  |  |  |  |  |
| I’ve been feeling useful (2) |  |  |  |  |  |
| I’ve been feeling relaxed (3) |  |  |  |  |  |
| I’ve been dealing with problems well (6) |  |  |  |  |  |
| I’ve been thinking clearly (7) |  |  |  |  |  |
| I’ve been feeling close to other people (9) |  |  |  |  |  |
| I’ve been able to make up my own mind about things (11) |  |  |  |  |  |

Q13 Have you ever been told by a doctor or nurse that you have depression or anxiety?

- Yes (5)
- No (6)
- Prefer not to answer (7)

End of Block: Physical and mental health

Start of Block: Fruit and vegetable consumption

Q22 Now we are moving on to a different topic, and we would like to ask you a few questions about some of the things you ate and drank yesterday. By yesterday we mean 24 hours from midnight to midnight.

Q23 Did you eat any salad yesterday? Don't count potato, pasta or rice salad or salad in a sandwich.

- Yes (47)
- No (48)

Display This Question:

If Did you eat any salad yesterday? Don't count potato, pasta or rice salad or salad in a sandwich. = Yes

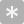

Q24 How many cereal bowls full of salad did you eat yesterday? (You can record half bowls of salad, such as 1.5, 0.5, etc.)

________________________________________________________________

Q35 Did you eat any pulses yesterday? By pulses we mean lentils and all kinds of peas and beans, including chickpeas and baked beans. Don't count pulses in foods like Chilli con carne.

- Yes (47)
- No (48)

Display This Question:

If Did you eat any pulses yesterday? By pulses we mean lentils and all kinds of peas and beans, incl... = Yes

Q36 How many tablespoons of pulses did you eat yesterday?

- Lentils (1) ________________________________________________
- Peas (2) ________________________________________________
- Baked beans (3) ________________________________________________
- Beans (9) ________________________________________________
- Chickpeas (4) ________________________________________________
- Other (5) ________________________________________________

Q37 Not counting potatoes, did you eat any vegetables yesterday? Please include fresh, raw, tinned and frozen vegetables.

- Yes (47)
- No (48)

Display This Question:

If Not counting potatoes, did you eat any vegetables yesterday? Please include fresh, raw, tinned an... = Yes

Q38 How many tablespoons of vegetables did you eat yesterday?

- Carrots (1) ________________________________________________
- Tomatoes (2) ________________________________________________
- Broccoli (3) ________________________________________________
- Peppers (9) ________________________________________________
- Corn (4) ________________________________________________
- Cabbage (5) ________________________________________________
- Other (10) ________________________________________________

Q39 Apart from anything you have already told us about, did you eat any other dishes made mainly from vegetables or pulses yesterday, such as vegetable lasagne or vegetable curry? Don't count vegetable soups or dishes made mainly from potatoes.

- Yes (47)
- No (48)

Display This Question:

If Apart from anything you have already told us about, did you eat any other dishes made mainly from... = Yes

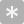

Q40 How many tablespoons of vegetables or pulses did you eat in these kinds of dishes yesterday?

________________________________________________________________

Q41 Compared with the amount of vegetables, salads and pulses you usually eat (when not in lockdown), would you say that yesterday you ate...

|  | Much less than usual (1) | Less than usual (2) | About the same as usual (3) | More than usual (4) | Much more than usual (5) |
| --- | --- | --- | --- | --- | --- |
| (1) |  |  |  |  |  |

Q42 Not counting cordials, fruit-drinks and squashes, did you drink any fruit juice yesterday?

- Yes (47)
- No (48)

Display This Question:

If Not counting cordials, fruit-drinks and squashes, did you drink any fruit juice yesterday? = Yes

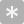

Q43 How many small glasses of fruit juice did you drink yesterday? A small glass is about a quarter of a pint.

________________________________________________________________

Q44 Did you eat any fresh fruit yesterday? Don't count fruit salads, fruit pies, etc.

- Yes (47)
- No (48)

Display This Question:

If Did you eat any fresh fruit yesterday? Don't count fruit salads, fruit pies, etc. = Yes

Q45 How many of these kinds of fresh fruit did you eat yesterday?

- Bananas (1) ________________________________________________
- Apples (2) ________________________________________________
- Oranges/satsumas/mandarins (3) ________________________________________________
- Grapes (handfuls) (9) ________________________________________________
- Other (4) ________________________________________________

Q46 Did you eat any dried fruit yesterday? Don't count dried fruit in cereal, cakes, etc.

- Yes (47)
- No (48)

Display This Question:

If Did you eat any dried fruit yesterday? Don't count dried fruit in cereal, cakes, etc. = Yes

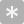

Q47 How many tablespoons of dried fruit did you eat yesterday?

________________________________________________________________

Q48 Did you eat any frozen fruit yesterday?

- Yes (47)
- No (48)

Display This Question:

If Did you eat any frozen fruit yesterday? = Yes

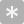

Q49 How many tablespoons of frozen fruit did you eat yesterday?

________________________________________________________________

Q50 Did you eat any tinned fruit yesterday?

- Yes (47)
- No (48)

Display This Question:

If Did you eat any tinned fruit yesterday? = Yes

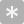

Q51 How many tablespoons of tinned fruit did you eat yesterday?

________________________________________________________________

Q52 Apart from anything you have already told us about, did you eat any other dishes made mainly from fruit yesterday, such as fruit salad or fruit pie? Don't count fruit in yoghurts.

- Yes (47)
- No (48)

Display This Question:

If Apart from anything you have already told us about, did you eat any other dishes made mainly from... = Yes

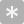

Q53 How many tablespoons of fruit did you eat in these kinds of dishes yesterday?

________________________________________________________________

Q54 Compared with the amount of fruit and fruit juice you usually eat and drink (when not in lockdown), would you say that yesterday you ate and drank...

|  | Much less than usual (1) | Less than usual (2) | About the same as usual (3) | More than usual (4) | Much more than usual (5) |
| --- | --- | --- | --- | --- | --- |
| (1) |  |  |  |  |  |

End of Block: Fruit and vegetable consumption

Start of Block: Demographics

Q25 Finally, we would like to ask some questions about you. If you prefer not to answer any of these questions, you can do so, but it would really help our research to have this information, which we will only use strictly anonymously.

Q26 What is your gender?

- Female (11)
- Male (12)
- Other, please describe if you wish: (13) ________________________________________________
- Prefer not to answer (14)

Q27 What is your age (in years)?

- I am (24) ________________________________________________
- Prefer not to answer (25)

Q28 How tall are you without shoes? You can answer this question in centimetres or in feet and inches.

- Answer in centimetres (1)
- Answers in feet and inches (2)
- Prefer not to answer (4)

Display This Question:

If How tall are you without shoes? You can answer this question in centimetres or in feet and inches. = Answer in centimetres

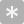

Q29 How tall are you without shoes? (in centimetres)

________________________________________________________________

Display This Question:

If How tall are you without shoes? You can answer this question in centimetres or in feet and inches. = Answers in feet and inches

Q30 How tall are you without shoes?

- Feet (4) ________________________________________________
- Inches (5) ________________________________________________

Q31 How much do you weigh without clothes and shoes? You can answer this question in kilograms or in stones and pounds.

- Answer in kilograms (1)
- Answer in stones and pounds (2)
- Prefer not to answer (4)

Display This Question:

If How much do you weigh without clothes and shoes? You can answer this question in kilograms or in... = Answer in kilograms

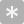

Q32 How much do you weigh without clothes and shoes? (in kilograms)

________________________________________________________________

Display This Question:

If How much do you weigh without clothes and shoes? You can answer this question in kilograms or in... = Answer in stones and pounds

Q33 How much do you weigh without clothes and shoes?

- Stones (5) ________________________________________________
- Pounds (6) ________________________________________________

Q34 Compared to the average household or family in the UK, would you say that your family is:

- Much poorer (1)
- Somewhat poorer (2)
- As rich (3)
- Slightly richer (4)
- Much richer (5)
- Prefer not to answer (6)

Q60 This was the last question. Please click the arrow below to complete the survey.

 If you are concerned about your mental health or wellbeing, [these helplines and support groups](https://www.nhs.uk/conditions/stress-anxiety-depression/mental-health-helplines/) can offer expert advice.

End of Block: Demographics
