## Supplement 1B. Questionnaire survey T1 for "Healthy diets, lifestyle changes and wellbeing during and after lockdown: Longitudinal evidence from the West Midlands"

Eating behaviour during Covid lockdown - Follow-up

Survey Flow

EmbeddedData

PROLIFIC_PIDValue will be set from Panel or URL.

Standard: Participant Information Sheet (2 Questions)

Branch: New Branch

If

If PARTICIPANT INFORMATION SHEET   Study Title: The impact of lockdown on health and wellbeing and f... I do not wish to participate Is Selected

EndSurvey: Advanced

Branch: New Branch

If

If PARTICIPANT INFORMATION SHEET   Study Title: The impact of lockdown on health and wellbeing and f... I have read the above and consent to take part in this study Is Selected

Block: Lockdown situation. (8 Questions)

Standard: Physical and mental health (5 Questions)

Standard: Fruit and vegetable consumption (25 Questions)

Standard: Demographics (9 Questions)

EndSurvey: Advanced

EndSurvey:

| Page Break |
| --- |

- I have read the above and consent to take part in this study (1)
- I do not wish to participate (4)

| 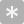 | 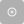 |
| --- | --- |

Q54 Please enter your Prolific ID:

________________________________________________________________

End of Block: Participant Information Sheet

Start of Block: Lockdown situation.

Q14 First, we would like to ask you some questions about the situation you are currently in. Here, we refer to the last two weeks, when the “lockdown” to control the Coronavirus had been largely released.

Q2 Are you currently

- Not working/studying (1)
- Working/studying from home (2)
- Working/studying at your workplace/school (3)
- Working/studying partly from home, partly at workplace/school (5)

Display This Question:

If Are you currently = Working/studying partly from home, partly at workplace/school

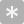

Q17 Roughly what percentage of the time are you working from home? (%)

________________________________________________________________

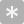

Q61 How many people are in your household, including yourself? Your household includes everyone living with you in the same house, sharing a kitchen.

________________________________________________________________

Display This Question:

If If How many people are in your household, including yourself? Your household includes everyone livin... Text Response Is Greater Than or Equal to 2

Q3 What ages are the members of your household, including yourself?

- Children 0-4 years (1) ________________________________________________
- Children 5-11 years (2) ________________________________________________
- Children 12-17 years (3) ________________________________________________
- Adults 18-64 years (12) ________________________________________________
- Adults 65-74 (13) ________________________________________________
- Adults 75+ (14) ________________________________________________

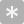

Q5 How many times per week do you exercise or go outdoors for active recreation (walk, sports, ...)?

________________________________________________________________

Q65 How do you usually get your shopping, and has this changed with the lockdown?

| Before the lockdown (1) | ▼ I use public transport (bus, taxi) to get my shopping (1) ... I rely on volunteers/others to get my shopping (6) |
| --- | --- |
| During the lockdown (6) | ▼ I use public transport (bus, taxi) to get my shopping (1) ... I rely on volunteers/others to get my shopping (6) |
| Now (after the lockdown) (7) | ▼ I use public transport (bus, taxi) to get my shopping (1) ... I rely on volunteers/others to get my shopping (6) |

Q9 When you are deciding what to buy when food shopping, how often do you consider your health?

|  | Never (1) | Seldom (3) | Sometimes (4) | Often (5) | Almost always (6) |
| --- | --- | --- | --- | --- | --- |
| (6) |  |  |  |  |  |

End of Block: Lockdown situation.

Start of Block: Physical and mental health

Q21 We would now like to ask you some questions about your health during the last two weeks, when the “lockdown” to control the Coronavirus had been largely released.

Q68 How was your health in general during the last 2 weeks?

|  | Very bad (1) | Bad (2) | Fair (3) | Good (4) | Very good (5) |
| --- | --- | --- | --- | --- | --- |
| (6) |  |  |  |  |  |

Q10 For each of the following four questions, please give an answer on a scale from 0 to 10, where 0 is "not at all" and 10 is "completely".

|  | 0 (not at all) (1) | 2.5 (6) | 5 (7) | 7.5 (9) | 10 (completely) (5) |
| --- | --- | --- | --- | --- | --- |
| All things considered, how satisfied were you with your life over the last 2 weeks? (1) |  |  |  |  |  |
| Overall, to what extent do you feel that the things that you do in your life are worthwhile? (6) |  |  |  |  |  |
| Overall, how happy did you feel yesterday? (7) |  |  |  |  |  |
| On a scale where 0 is "not at all anxious" and 10 is "completely anxious", overall, how anxious did you feel yesterday? (8) |  |  |  |  |  |

Q12 Which of these best describes your experience over the last 2 weeks?

|  | None of the time (1) | Rarely (2) | Some of the time (3) | Often (4) | All of the time (5) |
| --- | --- | --- | --- | --- | --- |
| I’ve been feeling optimistic about the future (1) |  |  |  |  |  |
| I’ve been feeling useful (2) |  |  |  |  |  |
| I’ve been feeling relaxed (3) |  |  |  |  |  |
| I’ve been dealing with problems well (6) |  |  |  |  |  |
| I’ve been thinking clearly (7) |  |  |  |  |  |
| I’ve been feeling close to other people (9) |  |  |  |  |  |
| I’ve been able to make up my own mind about things (11) |  |  |  |  |  |

Q11 Compared with how satisfied you usually were with your life (before the lockdown), would you say that you felt:

|  | Much less satisified than usual (1) | Less satisfied than usual (2) | About the same as usual (3) | More satisfied than usual (4) | Much more satisfied than usual (5) |
| --- | --- | --- | --- | --- | --- |
| During the lockdown (8) |  |  |  |  |  |
| Now (after the lockdown) (9) |  |  |  |  |  |

End of Block: Physical and mental health

Start of Block: Fruit and vegetable consumption

Q22 Now we are moving on to a different topic, and we would like to ask you a few questions about some of the things you ate and drank yesterday. By yesterday we mean 24 hours from midnight to midnight.

Q23 Did you eat any salad yesterday? Don't count potato, pasta or rice salad or salad in a sandwich.

- Yes (47)
- No (48)

Display This Question:

If Did you eat any salad yesterday? Don't count potato, pasta or rice salad or salad in a sandwich. = Yes

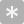

Q24 How many cereal bowls full of salad did you eat yesterday? (You can record half bowls of salad, such as 1.5, 0.5, etc.)

________________________________________________________________

Q35 Did you eat any pulses yesterday? By pulses we mean lentils and all kinds of peas and beans, including chickpeas and baked beans. Don't count pulses in foods like Chilli con carne.

- Yes (47)
- No (48)

Display This Question:

If Did you eat any pulses yesterday? By pulses we mean lentils and all kinds of peas and beans, incl... = Yes

Q36 How many tablespoons of pulses did you eat yesterday?

- Lentils (1) ________________________________________________
- Peas (2) ________________________________________________
- Baked beans (3) ________________________________________________
- Beans (9) ________________________________________________
- Chickpeas (4) ________________________________________________
- Other (5) ________________________________________________

Q37 Not counting potatoes, did you eat any vegetables yesterday? Please include fresh, raw, tinned and frozen vegetables.

- Yes (47)
- No (48)

Display This Question:

If Not counting potatoes, did you eat any vegetables yesterday? Please include fresh, raw, tinned an... = Yes

Q38 How many tablespoons of vegetables did you eat yesterday?

- Carrots (1) ________________________________________________
- Tomatoes (2) ________________________________________________
- Broccoli (3) ________________________________________________
- Peppers (9) ________________________________________________
- Corn (4) ________________________________________________
- Cabbage (5) ________________________________________________
- Other (10) ________________________________________________

Q39 Apart from anything you have already told us about, did you eat any other dishes made mainly from vegetables or pulses yesterday, such as vegetable lasagne or vegetable curry? Don't count vegetable soups or dishes made mainly from potatoes.

- Yes (47)
- No (48)

Display This Question:

If Apart from anything you have already told us about, did you eat any other dishes made mainly from... = Yes

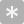

Q40 How many tablespoons of vegetables or pulses did you eat in these kinds of dishes yesterday?

________________________________________________________________

Q42 Not counting cordials, fruit-drinks and squashes, did you drink any fruit juice yesterday?

- Yes (47)
- No (48)

Display This Question:

If Not counting cordials, fruit-drinks and squashes, did you drink any fruit juice yesterday? = Yes

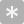

Q43 How many small glasses of fruit juice did you drink yesterday? A small glass is about a quarter of a pint.

________________________________________________________________

Q44 Did you eat any fresh fruit yesterday? Don't count fruit salads, fruit pies, etc.

- Yes (47)
- No (48)

Display This Question:

If Did you eat any fresh fruit yesterday? Don't count fruit salads, fruit pies, etc. = Yes

Q45 How many of these kinds of fresh fruit did you eat yesterday?

- Bananas (1) ________________________________________________
- Apples (2) ________________________________________________
- Oranges/satsumas/mandarins (3) ________________________________________________
- Grapes (handfuls) (9) ________________________________________________
- Other (4) ________________________________________________

Q46 Did you eat any dried fruit yesterday? Don't count dried fruit in cereal, cakes, etc.

- Yes (47)
- No (48)

Display This Question:

If Did you eat any dried fruit yesterday? Don't count dried fruit in cereal, cakes, etc. = Yes

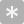

Q47 How many tablespoons of dried fruit did you eat yesterday?

________________________________________________________________

Q48 Did you eat any frozen fruit yesterday?

- Yes (47)
- No (48)

Display This Question:

If Did you eat any frozen fruit yesterday? = Yes

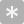

Q49 How many tablespoons of frozen fruit did you eat yesterday?

________________________________________________________________

Q50 Did you eat any tinned fruit yesterday?

- Yes (47)
- No (48)

Display This Question:

If Did you eat any tinned fruit yesterday? = Yes

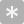

Q51 How many tablespoons of tinned fruit did you eat yesterday?

________________________________________________________________

Q52 Apart from anything you have already told us about, did you eat any other dishes made mainly from fruit yesterday, such as fruit salad or fruit pie? Don't count fruit in yoghurts.

- Yes (47)
- No (48)

Display This Question:

If Apart from anything you have already told us about, did you eat any other dishes made mainly from... = Yes

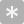

Q53 How many tablespoons of fruit did you eat in these kinds of dishes yesterday?

________________________________________________________________

Q73 Compared with the amount of fruit and vegetables you usually ate before the lockdown, would you say that you ate and drank...

|  | Much less (6) | Less (7) | About the same (8) | More (9) | Much more (10) |
| --- | --- | --- | --- | --- | --- |
| During the lockdown (8) |  |  |  |  |  |
| Now (after the lockdown) (9) |  |  |  |  |  |

Display This Question:

If Compared with the amount of fruit and vegetables you usually ate before the lockdown, would you s... != During the lockdown [ About the same ]

Q74 Can you please describe the reason(s) **why** your consumption of fruit and vegetables changed **during the lockdown**? (optional - you may leave this question blank)*[For instance: changes to shopping habits, motivation to eat healthily, less/more money available]*

________________________________________________________________

________________________________________________________________

________________________________________________________________

________________________________________________________________

________________________________________________________________

Display This Question:

If Compared with the amount of fruit and vegetables you usually ate before the lockdown, would you s... != Now (after the lockdown) [ About the same ]

Q75 Can you please describe the reason(s) **why** your consumption of fruit and vegetables is different **now**, compared to before the lockdown? (**optional - you may leave this question blank**)*[For instance: changes to shopping habits, motivation to eat healthily, less/more money available]*

________________________________________________________________

________________________________________________________________

________________________________________________________________

________________________________________________________________

________________________________________________________________

Q70 Do you agree or disagree with the following statements about your food shopping?

|  | Strongly agree (13) | Somewhat agree (14) | Neither agree nor disagree (15) | Somewhat disagree (16) | Strongly disagree (17) |
| --- | --- | --- | --- | --- | --- |
| I can afford to buy the fruit and vegetables I want in my usual shops (1) |  |  |  |  |  |
| Buying more fruit and vegetables would be difficult on my budget (2) |  |  |  |  |  |
| Lack of money prevents me from eating healthily (3) |  |  |  |  |  |

End of Block: Fruit and vegetable consumption

Start of Block: Demographics

Q25 Finally, we would like to ask some questions about you. If you prefer not to answer any of these questions, you can do so, but it would really help our research to have this information, which we will only use strictly anonymously.

Q31 How much do you weigh without clothes and shoes? You can answer this question in kilograms or in stones and pounds.

- Answer in kilograms (1)
- Answer in stones and pounds (2)
- Prefer not to answer (4)

Display This Question:

If How much do you weigh without clothes and shoes? You can answer this question in kilograms or in... = Answer in kilograms

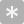

Q32 How much do you weigh without clothes and shoes? (in kilograms)

________________________________________________________________

Display This Question:

If How much do you weigh without clothes and shoes? You can answer this question in kilograms or in... = Answer in stones and pounds

Q33 How much do you weigh without clothes and shoes?

- Stones (5) ________________________________________________
- Pounds (6) ________________________________________________

Q76 How has your weight changed **compared to before the lockdown started**?

|  | I gained a lot of weight (6) | I gained some weight (7) | My weight is about the same (8) | I lost some weight (9) | I lost a lot of weight (10) |
| --- | --- | --- | --- | --- | --- |
| During the lockdown (8) |  |  |  |  |  |
| Now (after the lockdown) (9) |  |  |  |  |  |

Display This Question:

If How has your weight changed compared to before the lockdown started? != During the lockdown [ My weight is about the same ]

Or How has your weight changed compared to before the lockdown started? != Now (after the lockdown) [ My weight is about the same ]

Q77 What do you think is/are the reason(s) for the change in your weight? (**optional - you may leave this question blank**)*[For instance: changes to screen time, level of activity, social support and activities, sleeping, motivation to have a healthy lifestyle, eating behaviour, intake of alcohol, smoking]*

________________________________________________________________

________________________________________________________________

________________________________________________________________

________________________________________________________________

________________________________________________________________

Q62 What is your postcode area (first one or two letters of your postcode)?

▼ B-Birmingham (21) ... Prefer not to answer (16)

Display This Question:

If What is your postcode area (first one or two letters of your postcode)? != Other

And What is your postcode area (first one or two letters of your postcode)? != Prefer not to answer

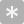

Q63 And the numbers of the **first part** of your postcode? (one or two digits only, please)

________________________________________________________________

Q60 This was the last question. Please click the arrow below to complete the survey.

 If you are concerned about your mental health or wellbeing, [these helplines and support groups](https://www.nhs.uk/conditions/stress-anxiety-depression/mental-health-helplines/) can offer expert advice.

End of Block: Demographics
