## Supplement 2. Distribution of fruit and vegetable consumption for "Healthy diets, lifestyle changes and wellbeing during and after lockdown: Longitudinal evidence from the West Midlands"

**Figure 3: Distribution of consumption of fruits and vegetables (portions/day) in the West Midlands**

HSE 2017, West Midlands:

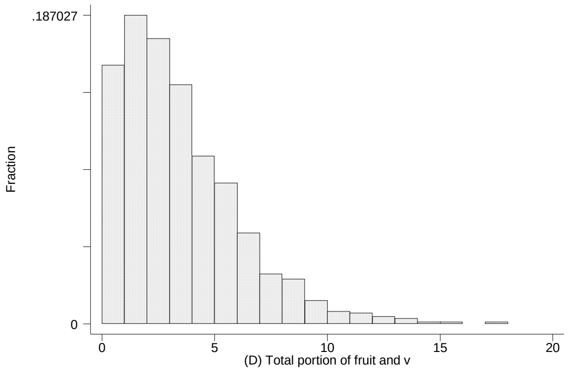


Mean: 3.2 portions/day, std.dev. 2.7 portions

Our sample:

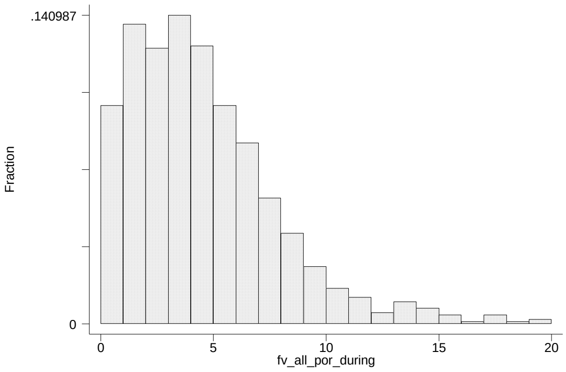


Mean: 4.6 portions/day, std.dev. 4.1 portions
