## Supplement 3. Heterogeneity in menal wellbeing for "Healthy diets, lifestyle changes and wellbeing during and after lockdown: Longitudinal evidence from the West Midlands"

### Supplement 3: Heterogeneity in mental wellbeing

#### Figure 4: Heterogeneity in the change (after versus during lockdown) in mental wellbeing (SWEMWBS)

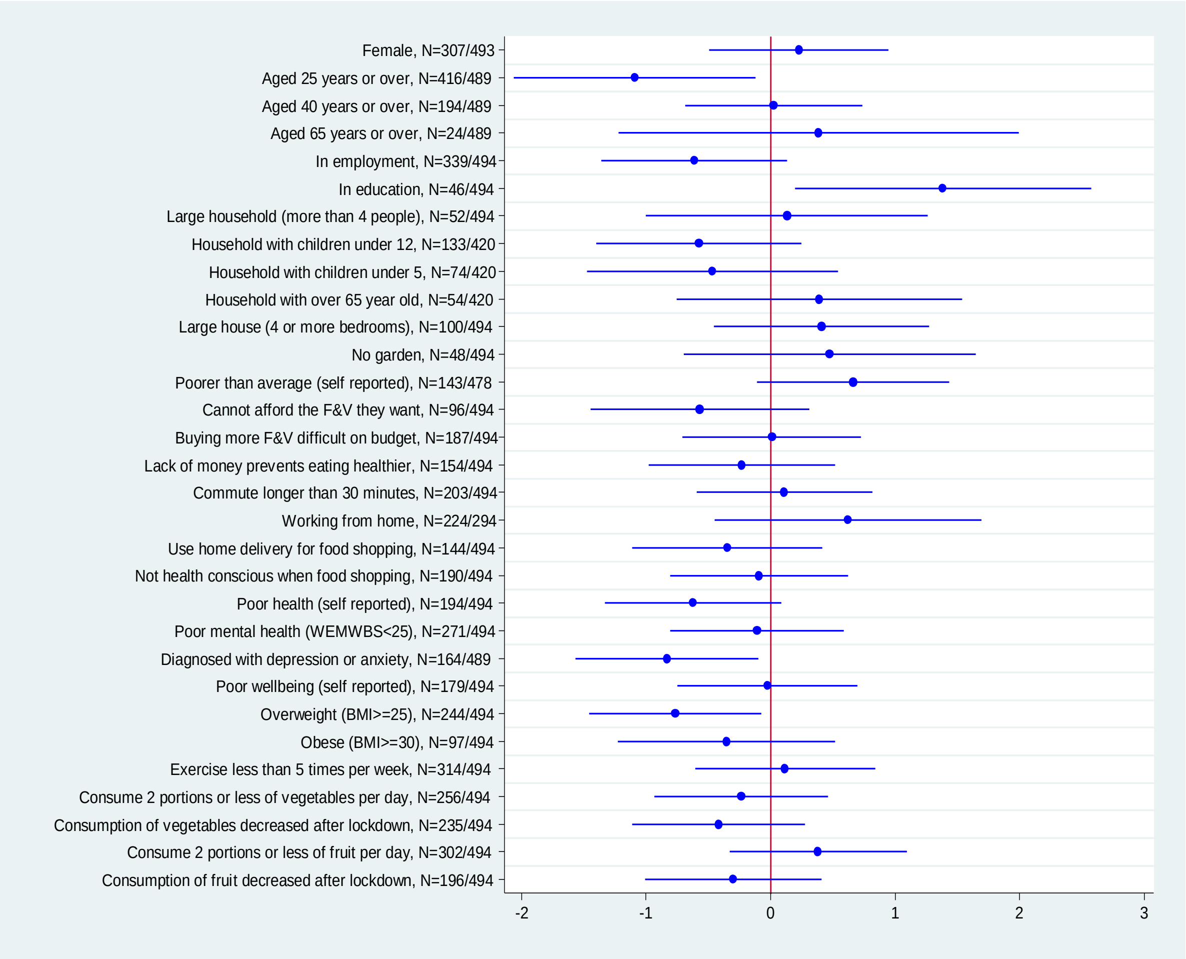
